## Appendix A for "Study protocol for the Innovative Support for Patients with SARS-COV-2 Infections Registry (INSPIRE): a longitudinal study of the medium and long-term sequelae of SARS-CoV-2 infection"

### INSPIRE

### Survey Specifications

Version 8.0
14 Dec 2021

### VERSION HISTORY

Detailed version history changes* available here.

| Draft/Version | Date | Changes Made |
|---|---|---|
| Version 1.0 | 10/20/20 | Final Draft |
| Version 1.1* | 12/09/20 | Updated language for dyspnea survey to remove "on the level" in response options |
| Version 2.0* | 01/06/21 | Updated screening question language |
| Version 3.0* | 03/02/21 | 1. Updated document formatting (cover page, page numbers, logo, question number)<br>2. Added Version History (pg. 2)<br>3. Added Table of Contents (pg. 3)<br>4. Added section headings<br>5. Added Appendix I: Instrument Sources (pg. 28)<br>6. Added Appendix II: Detailed Survey Version History (pg. 29)<br>7. Updated SDoH question schedule from "day 14" to "0 mon" (pg. 4)<br>8. Update Screening Questions (pg. 4)<br>9. Remove "on the level" from Dyspenea Questions (pg. 20) |
| Version 4.0* | 03/11/21 | 1. Added new variable types to Survey Summary and Schedule (pg. 4)<br>2. Added questions to 3 month follow up survey and quarterly follow up surveys (pg. 28):<br>a. Severity of illness<br>b. COVID-19 vaccination<br>c. Comorbidities<br>3. Updated Appendix II: Detailed Survey Version History (pg. 31) |
| Version 4.1* | 03/15/21 | Updated Spanish translations<br>1. Screening question 3<br>2. Testing question<br>3. PROMIS29 physical function question |
| Version 5.0 | 06/01/21 | 1. Updated Survey Summary Schedule Table with Survey Collection Type information ("FR" = Forced Response, "PNA" = Prefer Not to Answer)<br>2. Updated formatting of Baseline and Follow Up Survey questions to tables<br>3. Separated Follow Up Survey questions out from Baseline Survey questions<br>4. Indicated branching logic with brackets [→ SKIP TO NEXT] |
| Version 6.0* | 07/07/21 | 1. Updated Baseline and Follow Up Survey Questions for Symptom Assessment (Q4)<br>2. Added Baseline and Follow Up Survey Questions re: Visits to Healthcare Facilities |
| Version 6.1* | 07/15/21 | Updated screening symptom question from requiring at least 2 symptoms to 1 symptom (Q4)<br>Updated Spanish translation for screening symptom question instructions from select 2 to select at least 1 |
| Version 7.0* | 08/18/21 | Updated PROMIS 29 – Ability to Participate in Social Roles and Activities Domain answers to align with PROMIS-29 tool (pg. 26)<br>From: Not at all, Several Days, More than Half the Days, Nearly Every Day<br>To: Never, Rarely, Sometimes, Usually, Always |

| Draft/Version | Date | Changes Made |
|---|---|---|
| Version 8.0* | 12/14/21 | 1. Added “IRB Expectations for Changes to Survey” section (pg. 5)<br>2. Updated Table of Contents (pg. 6)<br><u>Screening Question Changes</u> (outlined in red)<br>3. Update formatting of Q3 (pg. 7):<br>4. Updated language of screening survey Q4 (pg. 7)<br><u>Baseline Survey Changes</u> (outlined in red)<br>5. Added “none of the above” option to Visit to Healthcare Facilities Q1 (pg.11)<br>6. Updated question type to FR for Visit to Healthcare Facilities Q1 (pg. 11)<br>7. Add language to Visit to Healthcare Facilities Q1 stem (pg. 11)<br>8. Remove “I never had and still do not have symptoms” option from Symptom Assessment Q1 responses (pg. 11)<br>9. Update the Symptom Assessment Q3 language (pg. 12)<br>10. Update the Symptom Assessment Q4 language (pg. 12)<br>11. Remove “living with a partner” response option for Marital Status Q4 (pg. 19)<br>12. Updated PROMIS29 Fatigue Q1 spelling (pg. 28)<br>13. Added vaccine survey questions to baseline survey (pg. 36)<br><u>Follow Up Survey Changes</u> (outlined in red)<br>14. Update Testing Q1 language (pg. 37)<br>15. Updated Testing Q2 responses (pg. 37)<br>16. Update Testing Q3 language (pg. 37)<br>17. Delete Testing Q4 (pg. 37)<br>18. Add “none of the above” option to Visit to Healthcare Facilities Q1 (pg.38)<br>19. Add language to Visit to Healthcare Facilities Q1 stem (pg. 38)<br>20. Remove Q1-3 from Symptom Assessment (pg. 38)<br>21. Update language of Symptom Assessment Q4 (pg. 38)<br>22. Add “None of the Above” to responses for Symptom Assessment Q4 (pg. 38)<br>23. Update Return to Work Q1 language (pg. 40)<br>24. Add “Not Applicable” response option to Return to Work Q1 (pg. 40)<br>25. Update Return to Work Q2 language (pg. 41)<br>26. Update Return to Work Q3 lanugage (pg. 41)<br>27. Update Return to Work Q3 responses (pg. 41)<br>28. Updated vaccine survey question language (pg. 43) |

#### IRB EXPECTATIONS FOR CHANGES TO SURVEY

This is a multi-site study with local IRB review and approval across 8 sites. The Clinical Core (at the UW) and the Executive Committee (Individuals from the Clinical Core, Analytic Core, Administrative Core, and the CDC) manage decisions on protocol-wide changes, including adjustments to the study surveys. Each time the survey is updated, however minor, IRB review and approval is required across all study sites before the survey can be updated in the online Hugo platform. As experienced so far, this effort across study sites and the Hugo platform, can take up to 3 months to accomplish one survey revision of minor changes. This creates a significant barrier in conducting this multi-site research, and a new, balanced approach is needed to move forward.

To increase the practicability to complete this CDC-funded research, this study is requesting IRB approval across all sites to build in the flexibility to make minor changes to the survey, without IRB modification, as outlined here:

- To make minor updates to the exact wording of existing IRB-approved questions and answers to improve language for respondents

To balance the needs of the study with the standards for IRB approval at the site level, each site is asked to further obtain IRB approval to allow for bulit-in approval to increase the practicability of the research to allow for minor survey updates during the conduct of the study, as outlined here:

- To add new questions that fall within the scope of the type of information currently outlined in the surveys (e.g., sociodemographics, clinical characteristics, symptom-related, testing information, vaccine-related, visits to healthcare, work-related, social determinants of health). New questions and responses will not increase risk to subjects (e.g., illegal drug use or overuse, behaviors that could place an individual at risk) or increase the identifiability of the study data (e.g., ask new identifiable information such as SSN).

### SURVEY SUMMARY AND SCHEDULE

| Variable Type | Instrument Source | Question Type | # Questions | Time Questions Asked | | | | | | | |
|---|---|---|---|---|---|---|---|---|---|---|---|
| | | | | 0 (Pre-Enrollment) | 0 (Baseline) | 3 | 6 | 9 | 12 | 15 | 18 |
| **Screening Questions** | INSPIRE Screening Criteria | | 4 | X | | | | | | | |
| **Sociodemographics (Stratification Questions)** | CDC PUI | | 4 | X | | | | | | | |
| **Testing information** | CDC PUI | | 3 | | X | | | | | | |
| **Visits to Healthcare Facilities** | | | 1 | | X | X | X | X | X | X | X |
| **Symptom check** | CDC PUI, case studies | | 23-44 | | X | X | X | X | X | X | X |
| **SDoH** | Multiple Sources | | 21 | | X | | | | | | |
| **Physical/mental health** | PROMIS-29 v2.1 | Forced Response | 29 | | X | X | X | X | X | X | X |
| **Cognitive Function** | PROMIS Cognitive SF 8a | Forced Response | 8 | | X | X | X | X | X | X | X |
| **Return to work/activity** | Global Health | Forced Response | 4 | | | X | X | X | X | X | X |
| **Post-infectious seq** | Dyspnea, cough | Forced Response | 6 | | X | X | X | X | X | X | X |
| **PTSD** | PC-PTSD-5 | Forced Response | 5 | | X | X | X | X | X | X | X |
| **Exercise** | Exercise vita sign | Forced Response | 2 | | X | X | X | X | X | X | X |
| **Fatigue symptoms** | CDC Short Symptom Screener | Forced Response | 20-103 | | X | X | X | X | X | X | X |
| **Severity of Illness** | | Forced Response | 1-2 | | | X | X | X | X | X | X |
| **Vaccine information** | | | 1-2 | | X | X | X | X | X | X | X |
| **Comorbidities** | | Forced Response | 1 | | | X | | | | | |
| **TOTAL** | | | **79-161** | | | | | | | | |

### PRE-ENROLLMENT SCREENING AND SOCIODEMOGRAPHIC QUESTIONS

| Variable Type | Instrument Source | # Questions | Time Questions Asked | | | | | | | |
|---|---|---|---|---|---|---|---|---|---|---|
| | | | 0 (Pre-Enrollment) | 0 (Baseline) | 3 | 6 | 9 | 12 | 15 | 18 |
| **Screening** | INSPIRE Screening Criteria | 4 | X | | | | | | | |
| **Sociodemographics** | CDC PUI | 4 | X | | | | | | | |

#### Screening Questions

| **Question Prompts** |
|---|
| These short questions will help determine your eligibility.<br>Next |
| Answer the following questions to see if you can take part<br>(Tap next to go on) |

| # | Type | Question | Response(s) |
|---|---|---|---|
| 1 | FR | Are you 18 years of age or older? | Yes/No |
| 2 | FR | Were you tested for COVID 19 in the last 42 days? (must be yes to be eligible) | Yes/No |
| 3 | FR | Have you tested positive for Covid-19 MORE THAN 42 DAYS ago? (if yes, ineligible) | Yes/No |
| 4 | FR | When your COVID-19 test was drawn, did you have any of the following symptoms at that time? (must select at least 1) | a. Fever<br>b. Chills<br>c. Myalgia (muscle aches)<br>d. Headache<br>e. Sore throat<br>f. Nausea or vomiting<br>g. Diarrhea<br>h. Fatigue (tiredness)<br>i. Congestion or runny nose<br>j. Cough<br>k. Shortness of breath<br>l. Difficulty breathing<br>m. New loss of taste or smell<br>n. None of the above |

#### Sociodemographic Characteristics

| # | Type | Questions | Answer(s) |
|---|---|---|---|
| 1 | PNA | What is your gender? | a. Male<br>b. Female<br>c. Transgender male<br>d. Transgender female<br>e. Non-binary/gender non-conforming<br>f. Not listed ________________<br>g. Prefer not to answer |
| 2 | PNA | Are you of Hispanic, Latin or Spanish Origin? | a. No, not of Hispanic, Latin or Spanish origin<br>b. Yes, Mexican/Mexican American<br>c. Yes, Puerto Rican<br>d. Yes, Cuban<br>e. Yes, another Hispanic, Latin or Spanish origin |
| 3 | PNA | Which category best describes your race? Check all that apply. [OMB/HHS Standards] | a. American Indian/Alaska Native<br>b. Asian [→SEE BRANCHING LOGIC]<br>c. Black or African American<br>d. Native Hawaiian/Other Pacific Islander [→SEE BRANCHING LOGIC]<br>e. White<br>f. Some other race (specify): ____________________ |
| | Skippable | | If Asian, please specify which sub-category (select as many as apply)<br><br>a. Chinese<br>b. Filipino<br>c. Asian Indian<br>d. Vietnamese<br>e. Korean<br>f. Japanese<br>g. Other Asian (e.g., Pakistani, Cambodian, Hmong) |
| | Skippable | | If Native Hawaiian/Other Pacific Islander, please specify sub-category (select as many as apply)<br><br>a. Native Hawaiian<br>b. Samoan<br>c. Chamorro<br>d. Other Pacific Islander (e.g., Tongan, Fijian, Mashallese) |

| | | | |
|---|---|---|---|
| 4 | PNA | What is the highest level of education you have completed? | a. 8th grade or less<br>b. Some high school, but did not graduate<br>c. High school graduate or GED<br>d. Some college but did not complete degree<br>e. 2-year college degree<br>f. 4-year college degree<br>g. More than 4-year college degree |

### BASELINE SURVEY QUESTIONS

#### Testing Information

| Variable Type | Instrument Source | # Questions | Time Questions Asked | | | | | | | |
|---|---|---|---|---|---|---|---|---|---|---|
| | | | 0 (Pre-Enrollment) | 0 (Baseline) | 3 | 6 | 9 | 12 | 15 | 18 |
| **Testing information** | CDC PUI | 3 | | X | See Follow Up Survey for follow up specific questions | | | | | |

| # | Type | Questions | Answer(s) |
|---|---|---|---|
| 1 | FR | Where did you last test for COVID? | a. Clinic including an Urgent Care Clinic<br>b. Tent/drive-up testing site<br>c. Emergency department<br>d. Hospital<br>e. At home testing kit<br>f. Other: _________ |
| 2 | FR | What was the approximate date of this test? | DATE [calendar format] |
| 3 | FR | Was the test positive for having COVID-19? | a. Yes<br>b. No<br>c. Waiting for results<br>d. Do not know |

#### Visits to Healthcare Facilities

| Variable Type | Instrument Source | # Questions | Time Questions Asked | | | | | | | |
|---|---|---|---|---|---|---|---|---|---|---|
| | | | 0 (Pre-Enrollment) | 0 (Baseline) | 3 | 6 | 9 | 12 | 15 | 18 |
| **Visits to Healthcare Facilities** | | 1 | | X | See Follow Up Survey for follow up specific questions | | | | | |

| # | Type | Questions | Answer(s) |
|---|---|---|---|
| 1 | FR | In the past month, have you: (Select as many that apply. If none apply, select "none of the above".) | a. Visited an outpatient clinic<br>b. Visited an urgent care center<br>c. Visited an emergency department<br>d. Been admitted overnight to hospital<br>e. Been admitted overnight to an intensive care unit/ward.<br>f. None of the above |

#### Symptom Assessment

| Variable Type | Instrument Source | # Questions | Time Questions Asked | | | | | | | |
|---|---|---|---|---|---|---|---|---|---|---|
| | | | 0 (Pre-Enrollment) | 0 (Baseline) | 3 | 6 | 9 | 12 | 15 | 18 |
| **Symptom check** | CDC PUI, case studies | 23-44 | | X | X | X | X | X | X | X |

| # | Type | Questions | Answer(s) |
|---|---|---|---|
| 1 | FR | Please give us your best estimate of when you first felt sick (including fever, cough, loss of smell or test, weakness, pain, etc.). | a. Date [calendar format] |
| 2 | FR | Give us your best estimate of when your symptoms ended. | a. DATE [calendar format]<br>I have ongoing symptoms now. |
| 3 | FR | On a scale of 1 to 10, with 1 being not so terrible and 10 being the worst, how bad did you feel at the peak of your illness/symptoms (when you felt the worst)? | Sliding bar |
| 4 | FR | Since you first felt sick with COVID-19 like symptoms, have you had any of the following (select yes for each)? | a. Fever >100.4F (38C)?<br>b. Feeling hot or feverish?<br>c. Chills?<br>d. Repeated shaking with chills?<br>e. More tired than usual?<br>f. Muscle aches?<br>g. Joint pains?<br>h. Runny nose<br>i. Sore throat?<br>j. A new cough, or worsening of a chronic cough?<br>k. Shortness of breath?<br>l. Wheezing?<br>m. Pain or tightness in your chest?<br>n. Palpitations?<br>o. Nausea or vomiting?<br>p. Headache?<br>q. Hair loss?<br>r. Abdominal pain?<br>s. Diarrhea (>3 loose/looser than normal stools/24 hours)?<br>t. Decreased smell or change in smell?<br>u. Decreased taste or change in taste?<br>v. Other (insert, specify which other sypmptoms as a write in text option) |

#### Fatigue Symptoms Questions

| Variable Type | Instrument Source | # Questions | Time Questions Asked | | | | | | | |
|---|---|---|---|---|---|---|---|---|---|---|
| | | | 0 (Pre-Enrollment) | 0 (Baseline) | 3 | 6 | 9 | 12 | 15 | 18 |
| **Fatigue symptoms** | CDC Short Symptom Screener | 20-103 | | X | X | X | X | X | X | X |

Below is a list of symptoms many individuals experience. Please tell us whether or not you have any of the symptoms listed below.
FOR EACH SYMPTOM, PLEASE CIRCLE THE APPROPRIATE ANSWERS IN THE GRID BELOW.

| | **DURING THE PAST MONTH, HAVE YOU HAD THIS SYMPTOM?** *PLEASE CIRCLE 1 OR 2. IF 1 IS SELECTED, THEN GO TO COLUMN A AND B* | | **COLUMN A** **PRIOR TO THIS PAST MONTH, FOR HOW LONG HAD YOU EXPERIENCED THIS SYMPTOM?** | | **COLUMN B** **DID YOU HAVE THIS SYMPTOM BEFORE THE RECENT HEALTH CONCERN THAT CAUSED YOU TO GET A COVID TEST?** | |
|---|---|---|---|---|---|---|
| | YES | NO | UNDER 6 MONTHS | 6 MONTHS OR LONGER | YES | NO |
| Fatigue, tiredness, or exhaustion | 1 | 2 | 1 | 2 | 1 | 2 |
| Muscle aches/muscle pains | 1 | 2 | 1 | 2 | 1 | 2 |
| Pain in joints | 1 | 2 | 1 | 2 | 1 | 2 |
| Unrefreshing sleep | 1 | 2 | 1 | 2 | 1 | 2 |
| Problems getting to sleep, sleeping through the night, or waking up on time | 1 | 2 | 1 | 2 | 1 | 2 |
| Forgetfulness/memory problems that caused you to substantially cut back on your activities | 1 | 2 | 1 | 2 | 1 | 2 |
| Difficulty thinking or concentrating that caused you to substantially cut back on your activities | 1 | 2 | 1 | 2 | 1 | 2 |
| Dizziness or fainting | 1 | 2 | 1 | 2 | 1 | 2 |

**1A.** For each symptom you noted in question 1, please fill in the grid to describe the frequency and the severity of the symptoms.

FOR EACH SYMPTOM, PLEASE CIRCLE THE APPROPRIATE ANSWERS IN THE GRID BELOW.

| SYMPTOMS SINTOMAS | DURING THE PAST MONTH, HOW OFTEN HAVE YOU HAD THIS SYMPTOM? | | | | | DURING THE PAST MONTH, HOW BAD WAS THIS SYMPTOM? | | | | |
|---|---|---|---|---|---|---|---|---|---|---|
| | A LITTLE OF THE TIME | SOME OF THE TIME | A GOOD BIT OF THE TIME | MOST OF THE TIME | ALL OF THE TIME | VERY MILD | MILD | MODERATE | SEVERE | VERY SEVERE |
| Fatigue, tiredness, or exhaustion | 1 | 2 | 3 | 4 | 5 | 1 | 2 | 3 | 4 | 5 |
| Muscle aches/muscle pains | 1 | 2 | 3 | 4 | 5 | 1 | 2 | 3 | 4 | 5 |
| Pain in joints | 1 | 2 | 3 | 4 | 5 | 1 | 2 | 3 | 4 | 5 |
| Unrefreshing sleep | 1 | 2 | 3 | 4 | 5 | 1 | 2 | 3 | 4 | 5 |
| Problems getting to sleep, sleeping through the night, or waking up on time | 1 | 2 | 3 | 4 | 5 | 1 | 2 | 3 | 4 | 5 |
| Forgetfulness/memory problems that caused you to substantially cut back on your activities | 1 | 2 | 3 | 4 | 5 | 1 | 2 | 3 | 4 | 5 |
| Difficulty thinking or concentrating that caused you to substantially cut back on your activities | 1 | 2 | 3 | 4 | 5 | 1 | 2 | 3 | 4 | 5 |
| Dizziness or fainting | 1 | 2 | 3 | 4 | 5 | 1 | 2 | 3 | 4 | 5 |

| # | Type | Questions | Answer(s) |
|---|---|---|---|
| **1B.a** | FR | [→IF FATIGUE, TIREDNESS, OR EXHAUSTION (ABOVE)] When this fatigue, tiredness, or exhaustion began, would you say that it came on all of a sudden, or slowly over time? | a. All of sudden<br>b. Slowly over time<br>c. Not aplicable<br>d. Don't know |
| **1B.b** | FR | [→IF FATIGUE, TIREDNESS, OR EXHAUSTION (ABOVE)] In what month and year did your fatiguing illness begin? | Month _____<br>Year_____ [dropdown list or calendar] |

| | | | |
|---|---|---|---|
| **1B.c** | FR | [→IF FATIGUE, TIREDNESS, OR EXHAUSTION (ABOVE)] When you are fatigued, does rest make your fatigue better? | 1 Yes, a lot<br>2 Yes, a little<br>3 No, not very much<br>6 Not applicable<br>8 Don't know |
| **1B.d** | FR | [→IF FATIGUE, TIREDNESS, OR EXHAUSTION (ABOVE)] When you are fatigued, has this fatigue substantially limited your ability to do occupational, educational, social, or personal activities? | 1 Yes<br>2 No<br>6 Not applicable<br>8 Don't know |
| **1C** | **FR** | Do any of them get worse for at least 24 hours after you engage in activities (physical or mental) that you were used to doing with no problems? | 1 Yes<br>2 No<br>6 Not applicable<br>8 Don't know |

#### Social Determinants Of Health Questions

| Variable Type | Instrument Source | # Questions | Time Questions Asked | | | | | | | |
|---|---|---|---|---|---|---|---|---|---|---|
| | | | 0 (Pre-Enrollment) | 0 (Baseline) | 3 | 6 | 9 | 12 | 15 | 18 |
| **SDoH** | | 21 | | X | | | | | | |

| Question Prompts |
|---|
| Sometimes social situations can affect your health and healing from being sick. We would like to ask you a few questions about this. |

| Household | | | |
|---|---|---|---|
| **#** | **Type** | **Questions** | **Answer(s)** |
| 1 | FR | Including yourself, how many adults aged 18-64 years are living in your household? | ENTER NUMERIC VALUE |
| 2 | FR | Including yourself, how many adults **over** the age of 65 are living in your household? | ENTER NUMERIC VALUE |
| 3 | FR | How many children (under the age of 18) are living in your household? | ENTER NUMERIC VALUE |

| Marital Status/Support (source: CMS/AHC) | | | |
|---|---|---|---|
| **#** | **Type** | **Questions** | **Answer(s)** |
| 4 | FR | What is your current marital status? | a. Married or civil union<br>b. Widowed<br>c. Divorced<br>d. Separated<br>e. Never married |
| **5** | FR | Is there someone around to help you if you need it (like taking you to the doctor, taking you grocery shopping, or making meals)? | a. Never<br>b. Rarely<br>*c.* Sometimes<br>d. Usually<br>e. Always |

| Housing (AHC HRSN Screening Tool; https://innovation.cms.gov/files/worksheets/ahcm-screeningtool.pdf) (source: CMS/AHC) | | | |
|---|---|---|---|
| **#** | **Type** | **Questions** | **Answer(s)** |
| 6 | FR | What is your living situation today? | a. I have a steady place to live<br>b. I have a place to live today, but I am worried about losing it in the future<br>c. I do not have a steady place to live (I am temporarily staying with others, in a hotel, in a shelter, living |

| | | | |
|---|---|---|---|
| | | | outside on the street, on a beach, in a car, abandoned building, bus or train station, or in a park |

| Income | | | |
|---|---|---|---|
| **#** | **Type** | **Questions** | **Answer(s)** |
| **7** | PNA | Prior to the COVID-19 pandemic, what was your annual household income from all sources? | a. Less than $10,000<br>b. $10,000 to $35,000<br>c. $35,000 to less than $50,000<br>d. $50,000 to less than $75,000<br>e. $75,000 or more |
| | FR | Has the COVID-19 pandemic caused you/your immediate family financial difficulties | a. Not at all<br>b. A little<br>c. Quite a bit<br>d. Very much |

| Food Security | | | |
|---|---|---|---|
| 8 | FR | In the past month, how often was the following true: "you worried that your food would run out before you got money to buy more." | a. Often true<br>b. Sometimes true<br>c. Never true |

| Utilities (CMS/AHC) | | | |
|---|---|---|---|
| **#** | **Type** | **Questions** | **Answer(s)** |
| **9** | PNA | In the past month, has the electric, gas, oil, or water company shut off services or threatened to shut off services (e.g., sending a warning notice) in your home? | a. No<br>b. Threatened to shut off services<br>c. Already shut off services |

| Transportation (CMS/AHC) | | | |
|---|---|---|---|
| **#** | **Type** | **Questions** | **Answer(s)** |
| 10 | FR | In the past month, has lack of reliable transportation kept you from medical appointments, meetings, work, or from getting to things needed for daily living? Check all that apply. | [CHECK ALL THAT APPLY]<br>a. Yes, it has kept me from medical appointments or getting medications<br>b. Yes, it has kept me from non-medical meetings, non-medical appointments, work, or getting things that I need<br>c. No |

| Access to Care *(NHANES/BRFSS/AHC CMS)* | | | |
|---|---|---|---|
| **#** | **Type** | **Questions** | **Answer(s)** |
| 11 | FR | Do you have any kind of health care coverage, including health insurance, prepaid plans such as HMOs, or government plans such as Medicaid, Medicare, or Indian Health Service? | CHECK ALL THAT APPLY<br>a. I do not have health insurance<br>b. State Children’s Health Insurance Plan<br>c. Medicare<br>d. Medicaid/State Insurance<br>e. Indian Health Service<br>f. Military<br>g. Private/Commercial/Employer-based/Self-insured |

| Employment: | | | |
|---|---|---|---|
| **#** | **Type** | **Questions** | **Answer(s)** |
| 12 | FR | Were you employed before the coronavirus outbreak? | a. Yes [→ SKIP QUESTION #13]<br>b. No [→ SKIP QUESTIONS #14 AND 15] |
| 13 | PNA | If not working, which best describes you? | *a.* Temporarily laid off, sick leave, or maternity leave<br>b. Looking for work, unemployed<br>c. Retired<br>d. Disabled, permanently, or temporarily<br>e. Keeping house<br>f. Student<br>g. Other (specify) |
| 14 | FR | Did you have a change in your job status since COVID? | a. No change<br>b. Reduced work hours<br>c. Permanently lost job<br>d. Temporarily lost job/Furloughed (kept job position, but was not paid for not working)<br>e. Increased hours |
| 15 | FR | Do you work in a healthcare setting such as a hospital, clinic, or nursing/rehabilitation care facility? | a. Yes<br>b. No [→ SKIP TO QUESTION #17] |

| | | | | |
|---|---|---|---|---|
| 16 | FR | What is your role in the hospital, clinic, nursing/rehabilitation care facility or other healthcare setting? | | *a.* Administrative (e.g., practice manager<br>b. Advanced Practice Provider<br>c. Chaplain/Religious Services<br>*d.* Clerk (e.g, registration clerk, unit clerk)<br>e. Environmental Services<br>f. Food<br>g. Laboratory Technician<br>h. Maintenance<br>i. Medical<br>j. Nurse<br>k. Nursing Assistant<br>l. Patient Care Technologist (Tech)<br>m. Pharmacist<br>n. Pharmacy Technician<br>o. Physician<br>p. Security<br>q. Transportation<br>r. Volunteer<br>s. Other ___________________ |
| 17 | FR | Are you a non-healthcare essential worker that was asked to work outside the home throughout the epidemic (e.g., police officer, bus driver, grocer, deliverer, manufacturer, store owner, chef/food supplier, teacher, etc). | | a. Yes<br>b. No |

| *Smoking/Alcohol/Prescription drugs/Illicit drugs* | | | | |
|---|---|---|---|---|
| **#** | **TYPE** | **Questions** | | **Answer(s)** |
| 18 | PNA | In the PAST 12 MONTHS, how often have you used tobacco or any other nicotine delivery product (i.e., e-cigarette, vaping or chewing tobacco)? | | a. Not at all<br>b. Daily or near daily<br>c. Weekly<br>d. Monthly<br>e. Less than monthly |
| 19 | PNA | In the PAST 12 MONTHS, how often have you had 5 or more drinks (men)/4 or more drinks (women) containing alcohol in one day? | | a. Not at all<br>b. Daily or near daily<br>c. Weekly<br>d. Monthly<br>e. Less than monthly |
| 20 | PNA | In the PAST 12 MONTHS, how often have you used any prescription medications just for the feeling, more than prescribed or that were not prescribed for you? | | a. Not at all<br>b. Daily or near daily<br>c. Weekly<br>d. Monthly<br>e. Less than monthly |
| 21 | PNA | In the PAST 12 MONTHS, how often have you smoked or vaped marijuana? | | a. Not at all<br>b. Daily or near daily<br>c. Weekly<br>d. Monthly<br>e. Less than monthly |
| 22 | PNA | In the PAST 12 MONTHS, how often have you used any drugs including cocaine or crack, heroin, methamphetamine (crystal meth), hallucinogens, ecstasy/MDMA? | | a. Not at all<br>b. Daily or near daily<br>c. Weekly<br>d. Monthly<br>e. Less than monthly |

#### PROMIS29 Questions

| Variable Type | Instrument Source | # Questions | Time Questions Asked | | | | | | | |
|---|---|---|---|---|---|---|---|---|---|---|
| | | | 0 (Pre-Enrollment) | 0 (Baseline) | 3 | 6 | 9 | 12 | 15 | 18 |
| **Physical/mental health** | PROMIS-29 v2.1 | 29 | | X | X | X | X | X | X | X |
| **Cognitive Function** | PROMIS Cognitive SF 8a | 8 | | X | X | X | X | X | X | X |

| ***(Promis29) Physical Function. In the last 7 days…*** | | | |
|---|---|---|---|
| **#** | **Type** | **Questions** | **Answer(s)** |
| 1 | FR | Were you able to do chores such as vacuuming or yard work? | a. Without any difficulty (5)<br>b. With a little difficulty (4<br>c. With some difficulty (3<br>d. With much difficulty (2)<br>e. Unable to do (1) |
| 2 | FR | Were you able to go up and down stairs at a normal pace? | a. Without any difficulty (5)<br>b. With a little difficulty (4<br>c. With some difficulty (3<br>d. With much difficulty (2)<br>e. Unable to do (1) |
| 3 | FR | Were you able to go for a walk of at least 15 minutes? | a. Without any difficulty (5)<br>b. With a little difficulty (4<br>c. With some difficulty (3<br>d. With much difficulty (2)<br>e. Unable to do (1) |
| 4 | FR | Were you able to run errands and shop? | a. Without any difficulty (5)<br>b. With a little difficulty (4<br>c. With some difficulty (3<br>d. With much difficulty (2)<br>e. Unable to do (1) |

| (Promis29) Anxiety. In the last 7 days… | | | |
|---|---|---|---|
| **#** | **Type** | **Questions** | **Answer(s)** |
| 5 | FR | I felt fearful. | a. Never (1)<br>b. Rarely (2)<br>*c.* Sometimes (3)<br>d. Often (4)<br>e. Always (5) |
| 6 | FR | I found it hard to focus on anything other than my anxiety | a. Never (1)<br>b. Rarely (2)<br>*c.* Sometimes (3)<br>d. Often (4)<br>e. Always (5) |

| | | | |
|---|---|---|---|
| 7 | FR | My worries overwhelmed me | a. Never (1)<br>b. Rarely (2)<br>*c.* Sometimes (3)<br>d. Often (4)<br>e. Always (5) |
| 8 | FR | I felt uneasy | a. Never (1)<br>b. Rarely (2)<br>*c.* Sometimes (3)<br>d. Often (4)<br>e. Always (5) |

| (Promis29) Depression. In the last 7 days… | | | |
|---|---|---|---|
| **#** | **Type** | **Questions** | **Answer(s)** |
| 9 | FR | I felt worthless | *a.* Never (1)<br>*b.* Rarely (2)<br>c. Sometimes (3)<br>d. Often (4)<br>e. Always (5) |
| 10 | FR | I felt helpless | *a.* Never (1)<br>*b.* Rarely (2)<br>c. Sometimes (3)<br>d. Often (4)<br>e. Always (5) |
| 11 | FR | I felt depressed | *a.* Never (1)<br>*b.* Rarely (2)<br>c. Sometimes (3)<br>d. Often (4)<br>e. Always (5) |
| 12 | FR | I felt hopeless | *a.* Never (1)<br>*b.* Rarely (2)<br>c. Sometimes (3)<br>d. Often (4)<br>e. Always (5) |

| (Promis29) Fatigue. In the last 7 days… | | | |
|---|---|---|---|
| **#** | **Type** | **Questions** | **Answer(s)** |
| 13 | FR | I feel fatigued | *a.* Never (1)<br>*b.* Rarely (2)<br>c. Sometimes (3)<br>d. Often (4)<br>e. Always (5) |
| 14 | FR | I have trouble starting things because I am tired | *a.* Never (1)<br>*b.* Rarely (2)<br>c. Sometimes (3)<br>d. Often (4)<br>e. Always (5) |

| | | | |
|---|---|---|---|
| 15 | FR | How run-down did you feel on average? | a. Not at all (1)<br>b. A little bit (2)<br>c. Somewhat (3)<br>d. Quite a bit (4)<br>e. Very much (5) |
| 16 | FR | How fatigued were you on average? | a. Not at all (1)<br>b. A little bit (2)<br>c. Somewhat (3)<br>d. Quite a bit (4)<br>e. Very much (5) |

| (Promis29) Ability to participate in social roles and activities | | | |
|---|---|---|---|
| **#** | **Type** | **Questions** | **Answer(s)** |
| 17 | FR | I have trouble doing all of my regular leisure activities with others | a. Never (5)<br>b. Rarely (4)<br>c. Sometimes (3)<br>d. Usually (2)<br>e. Always (1) |
| 18 | FR | I have trouble doing all of the family activities that I want to do | a. Never (5)<br>b. Rarely (4)<br>c. Sometimes (3)<br>d. Usually (2)<br>e. Always (1) |
| 19 | FR | I have trouble doing all of my usual work (include work at home). | a. Never (5)<br>b. Rarely (4)<br>c. Sometimes (3)<br>d. Usually (2)<br>e. Always (1) |
| 20 | FR | I have trouble doing all of the activities with friends that I want to do | a. Never (5)<br>b. Rarely (4)<br>c. Sometimes (3)<br>d. Usually (2)<br>e. Always (1) |

| (Promis29) Pain Interference. In the last 7 days | | | |
|---|---|---|---|
| **#** | **Type** | **Questions** | **Answer(s)** |
| 21 | FR | How much did pain interfere with your day to day activities? | a. Not at all (1)<br>b. A little bit (2)<br>c. Somewhat (3)<br>d. Quite a bit (4)<br>e. Very much (5) |
| 22 | FR | How much did pain interfere with work around the home? | a. Not at all (1)<br>b. A little bit (2)<br>c. Somewhat (3)<br>d. Quite a bit (4) |

| | | | |
|---|---|---|---|
| | | | e. Very much (5) |
| 23 | FR | How much did pain interfere with your ability to participate in social activities? | a. Not at all (1)<br>b. A little bit (2)<br>c. Somewhat (3)<br>d. Quite a bit (4)<br>e. Very much (5) |
| 24 | FR | How much did pain interfere with your household chores? | a. Not at all (1)<br>b. A little bit (2)<br>c. Somewhat (3)<br>d. Quite a bit (4)<br>e. Very much (5) |

| (Promis29) Pain Intensity. In the past 7 days | | | |
|---|---|---|---|
| **#** | **Type** | **Questions** | **Answer(s)** |
| 25 | FR | How would you rate your pain on average?. | Scale: No pain (0) to worst imaginable pain (10) |

| (Promis29) Sleep disturbance. In the past 7 days… | | | |
|---|---|---|---|
| **#** | **Type** | **Questions** | **Answer(s)** |
| 26 | FR | My sleep quality was | a. Very poor (5)<br>b. Poor (4)<br>c. Fair (3)<br>d. Good (2)<br>e. Very good (1) |
| 27 | FR | My sleep was refreshing | a. Not at all (1)<br>b. A little bit (2)<br>c. Somewhat (3)<br>d. Quite a bit (4)<br>e. Very much (5) |
| 28 | FR | I had a problem with my sleep | a. Not at all (1)<br>b. A little bit (2)<br>c. Somewhat (3)<br>d. Quite a bit (4)<br>e. Very much (5) |
| 29 | FR | I had difficulty falling asleep | a. Not at all (1)<br>b. A little bit (2)<br>c. Somewhat (3)<br>d. Quite a bit (4)<br>e. Very much (5) |

| (Promis Cognitive SF) Cognitive Function. In the past 7 days…. | | | |
|---|---|---|---|
| **#** | **Type** | **Questions** | **Answer(s)** |
| 1 | FR | My thinking has been slow | a. Never (5)<br>b. Rarely-Once (4)<br>c. Sometimes-Two or three times (3)<br>d. Often-About once a day (2) |

| | | | |
|---|---|---|---|
| | | | e. Very often-Several times a day (1) |
| 2 | FR | It has seemed like my brain was not working as well as usual | a. Never (5)<br>b. Rarely-Once (4)<br>c. Sometimes-Two or three times (3)<br>d. Often-About once a day (2)<br>e. Very often-Several times a day (1) |
| 3 | FR | I have had to work harder than usual to keep track of what I was doing | a. Never (5)<br>b. Rarely-Once (4)<br>c. Sometimes-Two or three times (3)<br>d. Often-About once a day (2)<br>e. Very often-Several times a day (1) |
| 4 | FR | I have had trouble shifting back and forth between different activities that require thinking | a. Never (5)<br>b. Rarely-Once (4)<br>c. Sometimes-Two or three times (3)<br>d. Often-About once a day (2)<br>e. Very often-Several times a day (1) |
| 5 | FR | I have had trouble concentrating | a. Never (5)<br>b. Rarely-Once (4)<br>c. Sometimes-Two or three times (3)<br>d. Often-About once a day (2)<br>e. Very often-Several times a day (1) |
| 6 | FR | I have had to work really hard to pay attention or I would make a mistake | a. Never (5)<br>b. Rarely-Once (4)<br>c. Sometimes-Two or three times (3)<br>d. Often-About once a day (2)<br>e. Very often-Several times a day (1) |
| 7 | FR | I have had trouble forming thoughts | a. Never (5)<br>b. Rarely-Once (4)<br>c. Sometimes-Two or three times (3)<br>d. Often-About once a day (2)<br>e. Very often-Several times a day (1) |
| 8 | FR | I have had trouble adding or subtracting numbers in my head | a. Never (5)<br>b. Rarely-Once (4)<br>c. Sometimes-Two or three times (3)<br>d. Often-About once a day (2)<br>e. Very often-Several times a day (1) |

#### Post Infectious Sequelae Questions

| Variable Type | Instrument Source | # Questions | Time Questions Asked | | | | | | | |
|---|---|---|---|---|---|---|---|---|---|---|
| | | | 0 (Pre-Enrollment) | 0 (Baseline) | 3 | 6 | 9 | 12 | 15 | 18 |
| **Post-infectious seq** | Dyspnea, cough | 6 | | X | X | X | X | X | X | X |

| (Modified Medical Research Council Dyspnea Scale) Dyspnea. | | | |
|---|---|---|---|
| # | Type | Questions | Answer(s) |
| 1 | FR | Please choose the one best response to describe your shortness of breath. | a. "I only get breathless with strenuous exercise" (0);<br>b. "I get short of breath when hurrying or walking up a slight hill" (1);<br>c. "I walk slower than people of the same age because of breathlessness or have to stop for breath when walking at my own pace" (2);<br>d. "I stop for breath after walking about 100 yards or after a few minutes" (3);<br>"I am too breathless to leave the house" or "I am breathless when dressing" (4) |

| (CCS) Cough symptom score system. Presently… | | | |
|---|---|---|---|
| # | Type | Questions | Answer(s) |
| 1 | FR | How frequently do you cough during the day? | a. None (1);<br>b. Seldom (2);<br>c. Sometimes (3);<br>d. Often (4);<br>e. All of the time (5) |
| 2 | FR | Does your cough disturb your sleep? | a. None (1);<br>b. Seldom (2);<br>c. Sometimes (3);<br>d. Often (4);<br>e. All of the time (5) |
| 3 | FR | Do you have an intense cough? | a. None (1);<br>b. Seldom (2);<br>c. Sometimes (3);<br>d. Often (4);<br>e. All of the time (5) |
| 4 | FR | Does your cough interfere with your daily life? | a. None (1);<br>b. Seldom (2);<br>c. Sometimes (3);<br>d. Often (4);<br>e. All of the time (5) |
| 5 | FR | Does your cough made you feel anxious or depressed? | a. None (1);<br>b. Seldom (2);<br>c. Sometimes (3);<br>d. Often (4);<br>e. All of the time (5) |

#### PTSD Questions

| Variable Type | Instrument Source | # Questions | Time Questions Asked | | | | | | | |
|---|---|---|---|---|---|---|---|---|---|---|
| | | | 0 (Pre-Enrollment) | 0 (Baseline) | 3 | 6 | 9 | 12 | 15 | 18 |
| **PTSD** | PC-PTSD-5 | 5 | | X | X | X | X | X | X | X |

| (PTSD5) PTSD. In the past month… | | | |
|---|---|---|---|
| **#** | **Type** | **Questions** | **Answer(s)** |
| 1 | FR | Had nightmares about the event(s) or thought about the event(s) when you did not want to? | a. Yes<br>b. No |
| 2 | FR | Tried hard not to think about the event(s) or went out of your way to avoid situations that reminded you of the event(s)? | a. Yes<br>b. No |
| 3 | FR | Been constantly on guard, watchful, or easily startled? | a. Yes<br>b. No |
| 4 | FR | Felt numb or detached from people, activities, or your surroundings? | a. Yes<br>b. No |
| 5 | FR | Felt guilty or unable to stop blaming yourself or others for the events(s) or any problems the event(s) may have caused? | a. Yes<br>b. No |

#### Exercise Questions

| Variable Type | Instrument Source | # Questions | Time Questions Asked | | | | | | | |
|---|---|---|---|---|---|---|---|---|---|---|
| | | | 0 (Pre-Enrollment) | 0 (Baseline) | 3 | 6 | 9 | 12 | 15 | 18 |
| **Exercise** | Exercise vital sign | 2 | | X | X | X | X | X | X | X |

| Exercise Vital Sign | | | |
|---|---|---|---|
| **#** | **Type** | **Questions** | **Answer(s)** |
| 1 | FR | For an average week in the last 30 days, how many days per week do you engage in moderate to strenuous exercise (like walking fast, running, jogging, dancing, swimming, biking, or other activities that cause a light or heavy sweat)? | Days/week [multiple choice] |
| 2 | FR | On those days that you engage in moderate to strenuous exercise, how many minutes, on average, do you exercise? | Minutes/day [forced numeric] |

#### Vaccine Related Questions

| Variable Type | Instrument Source | # Questions | Time Questions Asked | | | | | | | |
|---|---|---|---|---|---|---|---|---|---|---|
| | | | 0 (Pre-Enrollment) | 0 (Baseline) | 3 | 6 | 9 | 12 | 15 | 18 |
| **Vaccine information** | | 1-2 | | X | See Follow Up Survey for follow up specific questions | | | | | |

| | | | |
|---|---|---|---|
| **#** | **Type** | **Questions** | **Answer(s)** |
| 1 | Skippable | Have you had a COVID-19 vaccine? | a. Yes<br>b. No<br>c. Prefer not to answer |
| 1a | Skippable | [IF YES] To the best of your ability, when did you get vaccinated for COVID-19? | 1. Vaccine 1:<br>a. [CALENDAR – MONTH/YEAR]<br>b. Vaccine Type<br>i. Moderna<br>ii. Johnson & Johnson<br>iii. Pfizer<br>iv. Other<br>v. Do not know<br>2. Vaccine 2: (same as vaccine 1)<br>3. Vaccine 3: (same as vaccine 1) |

### FOLLOW UP SURVEY QUESTIONS

#### Testing Information

| Variable Type | Instrument Source | # Questions | Time Questions Asked | | | | | | | |
|---|---|---|---|---|---|---|---|---|---|---|
| | | | 0 (Pre-Enrollment) | 0 (Baseline) | 3 | 6 | 9 | 12 | 15 | 18 |
| **Testing information** | CDC PUI | 3 | | | X | X | X | X | X | X |

| # | Type | Questions | Answer(s) |
|---|---|---|---|
| 1 | FR | Since the last survey, have you had a positive COVID-19 test? | a. Yes<br>b. No [→ SKIP TO NEXT SECTION] |
| 2 | FR | If Yes....<br><br>Where was this study performed? | a. Clinic including an Urgent Care Clinic<br>b. Tent/drive-up testing site<br>c. Emergency department<br>d. Hospital<br>e. At home testing kit<br>f. Other: _________ |
| 3 | FR | What was the approximate date of this positive COVID-19 test? | DATE [calendar format] |

#### Visits to Healthcare Facilities

| Variable Type | Instrument Source | # Questions | Time Questions Asked | | | | | | | |
|---|---|---|---|---|---|---|---|---|---|---|
| | | | 0 (Pre-Enrollment) | 0 (Baseline) | 3 | 6 | 9 | 12 | 15 | 18 |
| **Visits to Healthcare Facilities** | | 1 | | | X | X | X | X | X | X |

| # | Type | Questions | Answer(s) |
|---|---|---|---|
| 1 | FR | Since you last told us about you and your health care, have you:<br>(Select as many that apply. If none apply, select None of the Above | a. Visited an out patient clinic<br>b. Visited an urgent care center<br>c. Visited an emergency department<br>d. Been admitted overnight to hospital<br>e. Been admitted overnight to an intensive care unit/ward.<br>f. None of the above |

#### Symptom Assessment

| Variable Type | Instrument Source | # Questions | Time Questions Asked | | | | | | | |
|---|---|---|---|---|---|---|---|---|---|---|
| | | | 0 (Pre-Enrollment) | 0 (Baseline) | 3 | 6 | 9 | 12 | 15 | 18 |
| **Symptom check** | CDC PUI, case studies | 1 | | | X | X | X | X | X | X |

| # | Type | Questions | Answer(s) |
|---|---|---|---|
| 1 | FR | Do you currently have any of the following ongoing symptoms? (Select as many that apply. If none apply, select None of the Above | a. Fever >100.4F (38C)?<br>b. Feeling hot or feverish?<br>c. Chills?<br>d. Repeated shaking with chills?<br>e. More tired than usual?<br>f. Muscle aches?<br>g. Joint pains?<br>h. Runny nose<br>i. Sore throat?<br>j. A new cough, or worsening of a chronic cough?<br>k. Shortness of breath?<br>l. Wheezing?<br>m. Pain or tightness in your chest?<br>n. Palpitations?<br>o. Nausea or vomiting?<br>p. Headache?<br>q. Hair loss?<br>r. Abdominal pain?<br>s. Diarrhea (>3 loose/looser than normal stools/24 hours)?<br>t. Decreased smell or change in smell?<br>u. Decreased taste or change in taste?<br>v. Other (insert, specify which other sypmptoms as a write in text option)<br>w. None of the Above |

#### Fatigue Symptoms Questions

| Variable Type | Instrument Source | # Questions | Time Questions Asked | | | | | | | |
|---|---|---|---|---|---|---|---|---|---|---|
| | | | 0 (Pre-Enrollment) | 0 (Baseline) | 3 | 6 | 9 | 12 | 15 | 18 |

| | | | | | | | | | | |
|---|---|---|---|---|---|---|---|---|---|---|
| **Fatigue symptoms** | CDC Short Symptom Screener | 20-103 | | X | X | X | X | X | X | X |

CLICK TO JUMP TO QUESTIONS

#### PROMIS29 Questions

| Variable Type | Instrument Source | # Questions | Time Questions Asked | | | | | | | |
|---|---|---|---|---|---|---|---|---|---|---|
| | | | 0 (Pre-Enrollment) | 0 (Baseline) | 3 | 6 | 9 | 12 | 15 | 18 |
| **Physical/mental health** | PROMIS-29 v2.: | 29 | | X | X | X | X | X | X | X |
| **Cognitive Function** | PROMIS Cognitive SF 8a | 8 | | X | X | X | X | X | X | X |

CLICK TO JUMP TO QUESTIONS

#### Return to Work Questions

| Variable Type | Instrument Source | # Questions | Time Questions Asked | | | | | | | |
|---|---|---|---|---|---|---|---|---|---|---|
| | | | 0 (Pre-Enrollment) | 0 (Baseline) | 3 | 6 | 9 | 12 | 15 | 18 |
| **Return to work/activity, global health** | | 4 | | | X | X | X | X | X | X |

| Return to work/activity and global health | | | |
|---|---|---|---|
| **#** | **Type** | **Questions** | **Answer (s)** |
| 1 | FR | [For people not retired, disabled, homemaker, not working/not looking for work] Did you return to work after your COVID-19 like symptoms? | a. Yes, full-time<br>b. Yes, part-time or modified work<br>c. No<br>d. Not applicable |
| 2 | FR | Since before you had COVID-19 like symptoms (if the first follow-up survey) or, since your last survey, how many workdays or weeks did you miss because of health reasons? | a. Don't work<br>b. 0-5 workdays<br>c. 6-10 workdays<br>d. 10-20 workdays<br>e. Up to 4 weeks |
| 3 | FR | Comparing your level of activity now with before you had COVID-19 like symptoms, would you say: | a. Your ability to do activities now is about the same as before you had COVID-19 like symptoms<br>b. Your ability to do activities now is somewhat less than before you had COVID-19 like symptoms<br>c. Your activity now is much less than before you had COVID-19 like symptoms |
| 4 | FR | How active are you during your leisure time at this point in time? | a. Mainly sedentary (e.g., sitting; reading; watching television)<br>b. Mild exercise (minimal effort; e.g., yoga; sport fishing; easy walking) |

| | | | |
|---|---|---|---|
| | | | c. Moderate exercise (e.g., walking; bicycle riding; gardening at least 4 hours a day)<br>d. Strenuous exercise (heart beats regularly, e.g., running/jogging; football; tennis; vigorous swimming) |
| 5 | FR | In general, how would you describe you health at this point in time? | a. 1 Excellent<br>b. 2 Very good<br>c. 3 Good<br>d. 4 Fair<br>e. 5 Poor<br>f. 98 Do not know<br>99 No answer |

#### Post Infectious Sequelae Questions

| Variable Type | Instrument Source | # Questions | Time Questions Asked | | | | | | | |
|---|---|---|---|---|---|---|---|---|---|---|
| | | | 0 (Pre-Enrollment) | 0 (Baseline) | 3 | 6 | 9 | 12 | 15 | 18 |
| **Post-infectious seq** | Dyspnea, cough | 6 | | X | X | X | X | X | X | X |

CLICK TO JUMP TO QUESTIONS

#### PTSD Questions

| Variable Type | Instrument Source | # Questions | Time Questions Asked | | | | | | | |
|---|---|---|---|---|---|---|---|---|---|---|
| | | | 0 (Pre-Enrollment) | 0 (Baseline) | 3 | 6 | 9 | 12 | 15 | 18 |
| **PTSD** | PC-PTSD-5 | 5 | | X | X | X | X | X | X | X |

CLICK TO JUMP TO QUESTIONS

#### Exercise Questions

| Variable Type | Instrument Source | # Questions | Time Questions Asked | | | | | | | |
|---|---|---|---|---|---|---|---|---|---|---|
| | | | 0 (Pre-Enrollment) | 0 (Baseline) | 3 | 6 | 9 | 12 | 15 | 18 |
| **Exercise** | Exercise vital sign | 2 | | X | X | X | X | X | X | X |

CLICK TO JUMP TO QUESTIONS

#### Severity of Illness Question

| | # Questions | Time Questions Asked |
|---|---|---|

| Variable Type | Instrument Source | | 0 (Pre-Enrollment) | 0 (Baseline) | 3 | 6 | 9 | 12 | 15 | 18 |
|---|---|---|---|---|---|---|---|---|---|---|
| **Severity of Illness** | | 1-2 | | | X | X | X | X | X | X |

| # | Type | Questions | Answer(s) |
|---|---|---|---|
| 1 | FR | Were you admitted to hospital for COVID-19? | a. Yes<br>b. No [→ SKIP TO NEXT SECTION] |
| 1a | FR | If Yes, in ICU? | a. Yes<br>No |

#### Vaccine Related Questions

| Variable Type | Instrument Source | # Questions | Time Questions Asked | | | | | | |
|---|---|---|---|---|---|---|---|---|---|
| | | | 0 (Pre-Enrollment) | 0 (Baseline) | 3 | 6 | 9 | 12 | 15 | 18 |
| **Vaccine information** | | 1-2 | | | X | X | X | X | X | X |

| # | Type | Questions | Answer(s) |
|---|---|---|---|
| 1 | Skippable | Have you had a COVID-19 vaccine? | a. Yes<br>b. No<br>c. Prefer not to answer |
| 1a | Skippable | [IF YES] To the best of your ability, when did you get vaccinated for COVID-19? | a. Have you had a COVID-19 vaccine?<br> a. Yes<br> b. No<br> c. Prefer not to answer<br>b. [IF YES] To the best of your ability, when did you get vaccinated for COVID-19?<br> a. I have already submitted this information and there are no changes to my vaccine status for COVID-19.<br> b. Vaccine 1:<br> i. [CALENDAR – MONTH/YEAR]<br> ii. Vaccine Type<br> 1. Moderna<br> 2. Johnson & Johnson<br> 3. Pfizer<br> 4. Other<br> 5. Do not know<br> c. Vaccine 2: (same as vaccine 1)<br> d. Vaccine 3: (same as vaccine 1) |

#### Comorbidity Question

| Variable Type | Instrument Source | # Questions | Time Questions Asked | | | | | | | |
|---|---|---|---|---|---|---|---|---|---|---|
| | | | 0 (Pre-Enrollment) | 0 (Baseline) | 3 | 6 | 9 | 12 | 15 | 18 |
| **Comorbidities** | | 1 | | | X | | | | | |

| # | Type | Questions | Answer(s) |
|---|---|---|---|
| 1 | FR | Do you have any of the following medical conditions (check all that apply) | a. Asthma (moderate or severe)<br>b. Hypertension or high blood pressure<br>c. Diabetes<br>d. Overweight orobesity<br>e. Emphysema or chronic obstructive pulmonary disease (COPD)<br>f. Heart conditions, such as coronary artery disease, heart failure or cardiomyopathies<br>g. Smoking (currently smoking any type of tobacco, including smokeless tobacco)<br>h. Kidney disease<br>i. Liver disease<br>j. Other (a text field will be added for write in's)<br>k. Don't know<br>l. Prefer not to answer |

### APPENDIX I: INSTRUMENT SOURCES

**INSTRUMENT SOURCES (at 0, 3, 6, 9, 12, 15, 18, 21 and 24 months)**
For each health domain, we will ask the initial questions listed in the "Instrument Source" column. When relevant, if they screen we will ask additional questions in the "Additional source" column.

| Health Domain | Instrument Source (No ques) | Number questions | Additional source |
|---|---|---|---|
| - Physical function<br>- Anxiety<br>- Depression<br>- Fatigue<br>- Sleep disturbance<br>- Ability to participate in social roles/activities | PROMIS SF v2.1- Physical Function 4a<br>PROMIS SFv1.0 – Anxiety 4a<br>PROMIS SF v1.0 – Depression 4a<br>PROMIS SF v.1.0- Sleep Disturbance 4a<br>PROMIS SF 1.0 – Ability to Particiapte in Social Roles and Activities 4a | 29 | See PROMIS-29v.2.1 PDF<br>On scoring the PROMIS-29: http://www.healthmeasures.net/images/PROMIS/manuals/PROMIS_Adult_Profile_Scoring_Manual.pdf |

| | | | |
|---|---|---|---|
| - Pain interference<br>- Pain intensity | PROMIS SF 1.0 Pain Interference 4a<br>PROMIS Pain Intensity Item (Global07) | | |
| Cognitive Function | PROMIS Cognitive Function-Short Form 8a<br>Administer if CDC Short Screener is positive | 8 | See PROMIS SF v. 2-Cognitive Function 8a pdf |
| Return to work/activity and global health | | 4 | Questions below |
| Dyspnea | Modified Medical Research Council Dyspnea Scale (1) | 1 | Question below |
| Cough | CCS: Cough Symptom Score System (5) | 5 | Questions below |
| PTSD | PC-PTSD-5 (5) | 5 | https://www.hiv.uw.edu/page/mental-health-screening/pc-ptsd |
| Exercise | Exercise Vital Sign (EVS) | 2 | https://pubmed.ncbi.nlm.nih.gov/22688832/ |
| Symptoms | CDC Short Symptom Screener | 2- 4 | In Section IV. Symptom Assessment above, see #6 |
| **TOTAL** | | **75-158** | |
